# Reevaluation of the Bicycle Ergometry in the Diagnosis of Ischemic Heart Disease

**DOI:** 10.1101/2024.07.03.24309879

**Authors:** Basheer Abdullah Marzoog, Peter Chomakhidze, Philipp Kopylov

## Abstract

**Background:** Ischemic heart disease (IHD) and related complications and outcomes remain the most frequently reported cardiovascular events. However, the diagnosis and prevention of ischemic heart disease primary depends on the clinical manifestations and electrocardiography changes.

**Objectives:** Patients with IHD are victims of the poor diagnostic methods and the prevention strategies.

**Aims:** To assess the diagnostic accuracy of the used bicycle ergometry in the diagnosis of ischemic heart disease based on the results of the stress computed myocardial perfusion (CTP) imaging with vasodilator test (adenosine triphosphate).

**Materials and methods:** A single center prospective, and non-randomized study included participations aged ≥ 40. The study included 38 participants with vs without IHD confirmed by CTP. The participants done bicycle ergometry following Bruce protocol then performed CTP. For statistical analysis used the descriptive statistics, Pearson Correlation test, Student test. The Statistica 12 program used for the statistical test.

**Results:** The study included 38 participants included 19 (50 %) patients with positive stress induced myocardial perfusion defect on the CTP and 19 (50 %) with negative results. The mean age of the participants 58,77 years (std. div. ± 9,664). The bicycle ergometry specificity and sensitivity 61.11%, 57.89%, respectively. The diagnostic accuracy of the bicycle ergometry 59.46%.

**Conclusions:** The bicycle ergometry test characterized by severe limitations in terms of the IHD diagnosis.

**Others:** Recommended to combine additional diagnostic methods with the bicycle ergometry to improve it is diagnostic accuracy, such as single channel electrocardiography and exhaled breath analysis.

## Introduction

Ischemic heart disease (IHD) is one of the most common pathologies among the cardiovascular disease in terms of the mortality and morbidity ^[1]^. This high prevalence of IHD in the community returns to the poor early detection and diagnostic methods of IHD. One of the most frequently used diagnostic method for individuals with clinical signs and symptoms of ischemic heart disease is the physical stress test, including bicycle ergometry ^[2]^. The debate about the efficacy of this method is under evaluation since it is first use in clinical practice ^[3, 4]^. However, this method is easy to use and safe as well as cost-effective. Therefore, this method is widespread as an elementary diagnostic method for IHD in compare to other expansive and invasive methods such as coronary angiography with Coronary Flow Reserve ^[5]^.

The current paper revises the specificity and sensitivity of the bicycle ergometry for usage as an elementary tool for IHD diagnosis through performing research on a participant with specific inclusion and exclusion criteria (published on the clinical trial (NCT06181799).

## Materials and methods

A non-randomized, single center, mini-invasive, parallel, cross-sectional, diagnostic, observational, case-control prospective cohort study performed at the University Clinical Hospital-1. The data of this study extracted from another clinical trial performed by Marzoog, the full design of the study published on clinicaltrials.gov (NCT06181799) ^[6]^. The study approved by the Sechenov University, Russia, from “Ethics Committee Requirement № 19-23 from 26.10.2023”.

The study included both males and females, and the age of the participants ≥ 40 years. The study participants passed exercise bicycle ergometry (on SCHILLER device; Bruce protocol) test to evaluate the response to physical activity. According to the results metabolic equivalent; Mets-BT (BT), the angina functional class (FC) in participants with positive physical stress test results determined, Where BT/Mets <50/<4 FC-III, BT/Mets 50-100/4-7 FC-II, BT/Mets >100/7 FC-I. During the bicycle ergometry, the participants monitored with 12-lead ECG and manual blood pressure measurement, 1 measured at the end of each 2 minute.

The ergometry procedure discontinued if an increase in blood pressure ≥ 220 mmHg or horizontal or downsloping ST segment on the ECG ≥ 1 mm. Moreover, stop the procedure if the target heart rate (86% of the 220-age) is reached.

To perform the stressed myocardial perfusion computer tomography imaging, all the participants present results of the venous creatinine level, eGFR (estimated glomerular filtration rate) according to the 2021 CKD-EPI Creatinine > 30 ml/min/1,73 m2, according to the recommendation for using this formula by the National kidney foundation and the American Society of Nephrology ^[7–10]^.

The participants got catheterization in the basilar vein or the radial vein for injection of contrast and Natrii adenosine triphosphate (10 mg/1ml) to induce pharmacological stress test to the heart by increasing heart rate.

To prepare the Natrii adenosine triphosphate, an Adenosine triphosphate 3 ml dilute in 17 ml of Isotonic Sodium Chloride Solution 0.9%, the injected volume of the diluted drug in milliliters is calculated by body weight. For 1 dose, take 3 ml of adenosine triphosphate (3 ampoules of each 1 ml (10mg)) + 17 ml of isotonic solution of sodium chloride 0.9% in one syringe, 20 ml. For one patient, manually inject intravenously (IV) through the already inserted catheter at a rate of 300 μg/kg/2 minutes, depending on weight: 60 kg = 12 ml, 70 kg = 14 ml. 80 kg = 16 ml, 100 kg = 20 ml of the full dose.

Computer tomography (Canon with 640 slice, 0,5mm thickness) with contrast (Omnipaque, 350-100ml), inject the contrast, then make rest image for myocardial perfusion, then wait 20 minutes and then inject the Natrii adenosine triphosphate intravenously (10 mg/1ml) according to the body weight to cause pharmacological stress test to the heart during two minutes in to the catheter. Then make the second image of the myocardial perfusion after stress test immediately.

## Results

The descriptive statistic of the continuous variables demonstrated in table 1.

**Table 1:** Descriptive statistics of the presented sample.

| Variable | Descriptive Statistics (All in primary data.stw) |  |  |  |  |
| --- | --- | --- | --- | --- | --- |
|  | Valid N | % Valid obs. | Mean | Minimum | Maximum |
| Age (years) | 38 | 100.0000 | 58.77 | 42.05 | 77.94 |
| Pulse rate (beat per minute) | 38 | 100.0000 | 72.21 | 53.00 | 92.00 |
| SBP (mmHg) | 38 | 100.0000 | 122.47 | 100.00 | 155.00 |
| DBP (mmHg) | 38 | 100.0000 | 77.84 | 60.00 | 100.00 |
| Body weight (Kg) | 38 | 100.0000 | 75.92 | 52.50 | 118.30 |
| Height (cm) | 38 | 100.0000 | 171.29 | 148.00 | 190.00 |
| BMI ($\text{kg/m}^2$) | 38 | 100.0000 | 25.84 | 18.49 | 35.00 |
| Goal heart rate ($220 - \text{age}$) | 38 | 100.0000 | 161.23 | 142.06 | 177.95 |
| Max HR (beat per minute) | 38 | 100.0000 | 148.08 | 108.00 | 199.00 |
| Reached % on bicycle ergometry | 38 | 100.0000 | 92.18 | 64.89 | 135.25 |
| WT | 38 | 100.0000 | 119.74 | 75.00 | 250.00 |
| METs | 38 | 100.0000 | 6.59 | 3.40 | 11.90 |
| Creatinine ($\mu\text{mol/L}$) | 38 | 100.0000 | 85.14 | 57.00 | 138.00 |
| eGFR (2021 CKD-EPI Creatinine) | 38 | 100.0000 | 82.04 | 45.40 | 107.00 |

The number of patients with normal body weight 17 (44.73684 %). overweight 15 (39.47368 %), and obesity first degree 6 (15.78947 %).

The categorical variables are presented as a relative and absolute values in table 2 (A-W)

**Table 2:** The table represents the categorical variables of the study in absolute and relative values.

| Category | Frequency table: Smoking (All in primary data.stw) |  |  |  |
| --- | --- | --- | --- | --- |
|  | Count | Cumulative Count | Percent | Cumulative Percent |
| No | 33 | 33 | 86.84211 | 86.8421 |
| Yes | 5 | 38 | 13.15789 | 100.0000 |
| Missing | 0 | 38 | 0.00000 | 100.0000 |

2-B
| Category | Frequency table: Concomitant disease (All in primary data.stw) |  |  |  |
| --- | --- | --- | --- | --- |
|  | Count | Cumulative Count | Percent | Cumulative Percent |
| No | 22 | 22 | 57.89474 | 57.8947 |
| Yes | 13 | 35 | 34.21053 | 92.1053 |
| Missing | 3 | 38 | 7.89474 | 100.0000 |

2-C
| Category | Frequency table: Atherosclerosis of the coronary artery (All in primary data.stw) |  |  |  |
| --- | --- | --- | --- | --- |
|  | Count | Cumulative Count | Percent | Cumulative Percent |
| No | 19 | 19 | 50.00000 | 50.0000 |
| Yes | 19 | 38 | 50.00000 | 100.0000 |
| Missing | 0 | 38 | 0.00000 | 100.0000 |

2-D
| Category | Frequency table: Hemodynamically significant (>60%) (All in primary data.stw) |  |  |  |
| --- | --- | --- | --- | --- |
|  | Count | Cumulative Count | Percent | Cumulative Percent |
| No | 33 | 33 | 86.84211 | 86.8421 |
| Yes | 5 | 38 | 13.15789 | 100.0000 |
| Missing | 0 | 38 | 0.00000 | 100.0000 |

2-E
| Category | Frequency table: Myocardial perfusion defect after stress ATP (All in primary data.stw) |  |  |  |
| --- | --- | --- | --- | --- |
|  | Count | Cumulative Count | Percent | Cumulative Percent |
| No | 19 | 19 | 50.00000 | 50.0000 |
|  | Count | Cumulative Count | Percent | Cumulative Percent |
| Yes | 19 | 38 | 50.00000 | 100.0000 |
| Missing | 0 | 38 | 0.00000 | 100.0000 |

2-G
| Category | Frequency table: Myocardial perfusion defect before Stress ATP (All in primary data.stw) |  |  |  |
| --- | --- | --- | --- | --- |
|  | Count | Cumulative Count | Percent | Cumulative Percent |
| Yes | 13 | 13 | 34.21053 | 34.2105 |
| No | 25 | 38 | 65.78947 | 100.0000 |
| Missing | 0 | 38 | 0.00000 | 100.0000 |

2-H
| Category | Frequency table: Atherosclerosis in other arteries (Yes/No) (All in primary data.stw) |  |  |  |
| --- | --- | --- | --- | --- |
|  | Count | Cumulative Count | Percent | Cumulative Percent |
| No | 18 | 18 | 47.36842 | 47.3684 |
| Yes | 15 | 33 | 39.47368 | 86.8421 |
| Missing | 5 | 38 | 13.15789 | 100.0000 |

2-I
| Category | Frequency table: Atherosclerotic vascular (Namely) (All in primary data.stw) |  |  |  |
| --- | --- | --- | --- | --- |
|  | Count | Cumulative Count | Percent | Cumulative Percent |
| Carotid, Brachiocephalic bifurcation | 12 | 12 | 31.57895 | 31.5789 |
| Carotid, Brachiocephalic bifurcation | 2 | 14 | 5.26316 | 36.8421 |
| Carotid | 1 | 15 | 2.63158 | 39.4737 |
| Missing | 23 | 38 | 60.52632 | 100.0000 |

2-J
| Category | Frequency table: Carotid (All in primary data.stw) |  |  |  |
| --- | --- | --- | --- | --- |
|  | Count | Cumulative Count | Percent | Cumulative Percent |
| No | 20 | 20 | 52.63158 | 52.6316 |
| Yes | 14 | 34 | 36.84211 | 89.4737 |
| Missing | 4 | 38 | 10.52632 | 100.0000 |

2-K
| Category | Frequency table: Brachiocephalic (All in primary data.stw) |  |  |  |
| --- | --- | --- | --- | --- |
|  | Count | Cumulative Count | Percent | Cumulative Percent |
| No | 22 | 22 | 57.89474 | 57.8947 |
|  | Count | Cumulative Count | Percent | Cumulative Percent |
| Yes | 12 | 34 | 31.57895 | 89.4737 |
| Missing | 4 | 38 | 10.52632 | 100.0000 |

2-L
| Category | Frequency table: Arterial Hypertension (All in primary data.stw) |  |  |  |
| --- | --- | --- | --- | --- |
|  | Count | Cumulative Count | Percent | Cumulative Percent |
| Yes | 21 | 21 | 55.26316 | 55.2632 |
| No | 17 | 38 | 44.73684 | 100.0000 |
| Missing | 0 | 38 | 0.00000 | 100.0000 |

2-M
| Category | Frequency table: Stage AH (All in primary data.stw) |  |  |  |
| --- | --- | --- | --- | --- |
|  | Count | Cumulative Count | Percent | Cumulative Percent |
| II | 14 | 14 | 36.84211 | 36.8421 |
| I | 2 | 16 | 5.26316 | 42.1053 |
| III | 5 | 21 | 13.15789 | 55.2632 |
| I | 1 | 22 | 2.63158 | 57.8947 |
| Missing | 16 | 38 | 42.10526 | 100.0000 |

2-N
| Category | Frequency table: Degree AH (All in primary data.stw) |  |  |  |
| --- | --- | --- | --- | --- |
|  | Count | Cumulative Count | Percent | Cumulative Percent |
| III | 5 | 5 | 13.15789 | 13.1579 |
| II | 5 | 10 | 13.15789 | 26.3158 |
| I | 11 | 21 | 28.94737 | 55.2632 |
| II | 1 | 22 | 2.63158 | 57.8947 |
| Missing | 16 | 38 | 42.10526 | 100.0000 |

2-O
| Category | Frequency table: Risk of CVD (All in primary data.stw) |  |  |  |
| --- | --- | --- | --- | --- |
|  | Count | Cumulative Count | Percent | Cumulative Percent |
| Moderate | 14 | 14 | 36.84211 | 36.8421 |
| High | 10 | 24 | 26.31579 | 63.1579 |
| Low | 11 | 35 | 28.94737 | 92.1053 |
| Very high | 3 | 38 | 7.89474 | 100.0000 |
| Missing | 0 | 38 | 0.00000 | 100.0000 |

2-P
| Category | Frequency table: SCAD (All in primary data.stw) |  |  |  |
| --- | --- | --- | --- | --- |
|  | Count | Cumulative Count | Percent | Cumulative Percent |
|  | Count | Cumulative Count | Percent | Cumulative Percent |
| No | 18 | 18 | 47.36842 | 47.3684 |
| Yes | 3 | 21 | 7.89474 | 55.2632 |
| Missing | 17 | 38 | 44.73684 | 100.0000 |

2-Q
| Category | Frequency table: Blood pressure reaction type (All in primary data.stw) |  |  |  |
| --- | --- | --- | --- | --- |
|  | Count | Cumulative Count | Percent | Cumulative Percent |
| Normotonic | 28 | 28 | 73.68421 | 73.6842 |
| Mild hypertonic | 2 | 30 | 5.26316 | 78.9474 |
| Hypertonic | 2 | 32 | 5.26316 | 84.2105 |
| Asthenic | 3 | 35 | 7.89474 | 92.1053 |
| Hypotonic | 3 | 38 | 7.89474 | 100.0000 |
| Missing | 0 | 38 | 0.00000 | 100.0000 |

2-R
| Category | Frequency table: FC by WT (All in primary data.stw) |  |  |  |
| --- | --- | --- | --- | --- |
|  | Count | Cumulative Count | Percent | Cumulative Percent |
| FC-II | 7 | 7 | 18.42105 | 18.4211 |
| FC-I | 5 | 12 | 13.15789 | 31.5789 |
| Missing | 26 | 38 | 68.42105 | 100.0000 |

2-S
| Category | Frequency table: FC by METs (All in primary data.stw) |  |  |  |
| --- | --- | --- | --- | --- |
|  | Count | Cumulative Count | Percent | Cumulative Percent |
| FC-II | 7 | 7 | 18.42105 | 18.4211 |
| FC-I | 4 | 11 | 10.52632 | 28.9474 |
| FC-III | 1 | 12 | 2.63158 | 31.5789 |
| Missing | 26 | 38 | 68.42105 | 100.0000 |

2-T
| Category | Frequency table: Reaction type ( positive/negative) (All in primary data.stw) |  |  |  |
| --- | --- | --- | --- | --- |
|  | Count | Cumulative Count | Percent | Cumulative Percent |
| Positive | 12 | 12 | 31.57895 | 31.5789 |
| Negative | 17 | 29 | 44.73684 | 76.3158 |
| Suspected | 9 | 38 | 23.68421 | 100.0000 |
| Missing | 0 | 38 | 0.00000 | 100.0000 |

2-U
| Category | Frequency table: Reason of discontinuation (All in primary data.stw) |  |  |  |
| --- | --- | --- | --- | --- |
|  | Count | Cumulative Count | Percent | Cumulative Percent |
| Horizontal ST depression >1mm | 6 | 6 | 15.78947 | 15.7895 |
| Reach goal HR | 32 | 38 | 84.21053 | 100.0000 |
| Missing | 0 | 38 | 0.00000 | 100.0000 |

2-V
| Category | Frequency table: Tolerance to exertion (All in primary data.stw) |  |  |  |
| --- | --- | --- | --- | --- |
|  | Count | Cumulative Count | Percent | Cumulative Percent |
| Close to high | 4 | 4 | 10.52632 | 10.5263 |
| Moderate | 23 | 27 | 60.52632 | 71.0526 |
| Very high | 6 | 33 | 15.78947 | 86.8421 |
| High | 4 | 37 | 10.52632 | 97.3684 |
| Low | 1 | 38 | 2.63158 | 100.0000 |
| Missing | 0 | 38 | 0.00000 | 100.0000 |

2-W
| Category | Frequency table: CKD stage (All in primary data.stw) |  |  |  |
| --- | --- | --- | --- | --- |
|  | Count | Cumulative Count | Percent | Cumulative Percent |
| I | 16 | 16 | 42.10526 | 42.1053 |
| II | 18 | 34 | 47.36842 | 89.4737 |
| IIIa | 4 | 38 | 10.52632 | 100.0000 |
| Missing | 0 | 38 | 0.00000 | 100.0000 |

The results of the student test of the sample represented in table 3 (A-M).

**Table 3:** The results of the student test of the sample represented. Only statistically significant difference represented in the table.

| Variable | T-tests; Grouping: Gender (All in primary data.stw) |  |  |  |  |  |
| --- | --- | --- | --- | --- | --- | --- |
|  | Group 1: F<br>Group 2: M |  |  |  |  |  |
|  | Mean F | Mean M | t-value | df | p | Valid N F |
| SBP rest | 117.41 | 126.57 | -2.40957 | 36 | 0.021210 | 17 |
| Body weight | 67.85 | 82.44 | -3.58890 | 36 | 0.000981 | 17 |
| Height | 164.62 | 176.69 | -4.83464 | 36 | 0.000025 | 17 |
| WT | 88.24 | 145.24 | -4.82673 | 36 | 0.000025 | 17 |
| METs | 5.81 | 7.22 | -2.24874 | 36 | 0.030743 | 17 |
| Creatinine (μmol/L) | 76.05 | 92.50 | -3.65241 | 36 | 0.000820 | 17 |

| Variable | T-tests; Grouping: Gender (All in primary data.stw) |
| --- | --- |
|  | Group 1: F<br>Group 2: M |

|  | Valid N<br>M | Std.Dev.<br>F | Std.Dev.<br>M | F-ratio<br>Variances | p<br>Variances |
| --- | --- | --- | --- | --- | --- |
| SBP rest | 21 | 12.654 | 10.782 | 1.377398 | 0.492763 |
| Body weight | 21 | 10.219 | 13.997 | 1.876169 | 0.205644 |
| Height | 21 | 8.795 | 6.600 | 1.775870 | 0.223898 |
| WT | 21 | 17.936 | 45.839 | 6.531701 | 0.000392 |
| METs | 21 | 1.206 | 2.358 | 3.822979 | 0.008835 |
| Creatinine (μmol/L) | 21 | 11.782 | 15.230 | 1.670764 | 0.301144 |

3-B
| Variable | T-tests; Grouping: Concomitant disease (All in primary data.stw)<br>Group 1: No<br>Group 2: Yes |  |  |  |  |  |
| --- | --- | --- | --- | --- | --- | --- |
|  | Mean<br>No | Mean<br>Yes | t-value | df | p | Valid N<br>No |
| Height | 173.27 | 165.62 | 2.52705 | 33 | 0.016476 | 22 |
| Reached % | 88.58 | 97.49 | -2.30415 | 33 | 0.027648 | 22 |
| WT | 130.68 | 98.08 | 2.13328 | 33 | 0.040428 | 22 |

| Variable | T-tests; Grouping: Concomitant disease (All in primary data.stw)<br>Group 1: No<br>Group 2: Yes |  |  |  |  |
| --- | --- | --- | --- | --- | --- |
|  | Valid N<br>Yes | Std.Dev.<br>No | Std.Dev.<br>Yes | F-ratio<br>Variances | p<br>Variances |
| Height | 13 | 8.062 | 9.622 | 1.424600 | 0.461098 |
| Reached % | 13 | 8.428 | 14.566 | 2.987445 | 0.027287 |
| WT | 13 | 52.288 | 21.558 | 5.882893 | 0.002912 |

3-C
| Variable | T-tests; Grouping: Atherosclerosis of the coronary artery (All in primary data.stw)<br>Group 1: No<br>Group 2: Yes |  |  |  |  |  |
| --- | --- | --- | --- | --- | --- | --- |
|  | Mean<br>No | Mean<br>Yes | t-value | df | p | Valid N<br>No |
| Age | 53.13 | 64.41 | -4.39659 | 36 | 0.000093 | 19 |
| Goal heart rate | 166.87 | 155.59 | 4.39659 | 36 | 0.000093 | 19 |
| Creatinine (μmol/L) | 79.40 | 90.89 | -2.35327 | 36 | 0.024191 | 19 |

| Variable | T-tests; Grouping: Atherosclerosis of the coronary artery (All in primary data.stw)<br>Group 1: No<br>Group 2: Yes |  |  |  |  |
| --- | --- | --- | --- | --- | --- |
|  | Valid N<br>Yes | Std.Dev.<br>No | Std.Dev.<br>Yes | F-ratio<br>Variances | p<br>Variances |
| Age | 19 | 6.825 | 8.850 | 1.68168 | 0.279471 |
| Goal heart rate | 19 | 6.825 | 8.850 | 1.68168 | 0.279471 |
| Creatinine (μmol/L) | 19 | 12.642 | 17.114 | 1.83250 | 0.208473 |

3-C
| Variable | T-tests; Grouping: Hemodynamically significant (>60%) (All in primary data.stw)<br>Group 1: No<br>Group 2: Yes |  |  |  |  |  |
| --- | --- | --- | --- | --- | --- | --- |
|  | Mean No | Mean Yes | t-value | df | p | Valid N No |
| SBP rest | 120.36 | 136.40 | -2.96948 | 36 | 0.005282 | 33 |
| Max HR | 150.79 | 130.20 | 2.97336 | 36 | 0.005229 | 33 |

| Variable | T-tests; Grouping: Hemodynamically significant (>60%) (All in primary data.stw)<br>Group 1: No<br>Group 2: Yes |  |  |  |  |
| --- | --- | --- | --- | --- | --- |
|  | Valid N Yes | Std.Dev. No | Std.Dev. Yes | F-ratio Variances | p Variances |
| SBP rest | 5 | 10.911 | 13.686 | 1.573274 | 0.410528 |
| Max HR | 5 | 13.713 | 19.215 | 1.963335 | 0.247939 |

3-E
| Variable | T-tests; Grouping: Myocardial perfusion defect before Stress ATP (All in primary data.stw)<br>Group 1: Yes<br>Group 2: No |  |  |  |  |  |
| --- | --- | --- | --- | --- | --- | --- |
|  | Mean Yes | Mean No | t-value | df | p | Valid N Yes |
| Max HR | 139.15 | 152.72 | -2.70213 | 36 | 0.010442 | 13 |
| Reached % | 86.04 | 95.37 | -2.46004 | 36 | 0.018825 | 13 |

| Variable | T-tests; Grouping: Myocardial perfusion defect before Stress ATP (All in primary data.stw)<br>Group 1: Yes<br>Group 2: No |  |  |  |  |
| --- | --- | --- | --- | --- | --- |
|  | Valid N No | Std.Dev. Yes | Std.Dev. No | F-ratio Variances | p Variances |
| Max HR | 25 | 14.899 | 14.573 | 1.045192 | 0.886020 |
| Reached % | 25 | 9.231 | 11.919 | 1.667134 | 0.357847 |

3-F
| Variable | T-tests; Grouping: Atherosclerosis in other arteries (Yes/No) (All in primary data.stw)<br>Group 1: No<br>Group 2: Yes |  |  |  |  |  |
| --- | --- | --- | --- | --- | --- | --- |
|  | Mean No | Mean Yes | t-value | df | p | Valid N No |
| Age | 52.16 | 65.61 | -5.29700 | 31 | 0.000009 | 18 |
| Goal heart rate | 167.84 | 154.39 | 5.29700 | 31 | 0.000009 | 18 |
| METs | 7.61 | 5.63 | 3.03459 | 31 | 0.004844 | 18 |

| Variable | T-tests; Grouping: Atherosclerosis in other arteries (Yes/No) (All in primary data.stw)<br>Group 1: No<br>Group 2: Yes |  |  |  |  |
| --- | --- | --- | --- | --- | --- |
|  | Valid N Yes | Std.Dev. No | Std.Dev. Yes | F-ratio Variances | p Variances |
| Age | 15 | 6.804 | 7.776 | 1.30590 | 0.594317 |
| Goal heart rate | 15 | 6.804 | 7.776 | 1.30590 | 0.594317 |
|  | Valid N<br>Yes | Std.Dev.<br>No | Std.Dev.<br>Yes | F-ratio<br>Variances | p<br>Variances |
| METs | 15 | 2.452 | 0.635 | 14.89954 | 0.000007 |

3-G
| Variable | T-tests; Grouping: Atherosclerotic vascular (Namely) (All in primary data.stw)<br>Group 1: Carotid, Brachiocephalic bifurcation<br>Group 2: Carotid, Brachiocephalic bifurcation |  |
| --- | --- | --- |
|  | Mean<br>Carotid, Brachiocephalic<br>bifurcation | Mean<br>Carotid, Brachiocephalic<br>bifurcation |
| Max HR | 139.33 | 184.00 |
| Reached % | 90.37 | 124.61 |

| Variable | T-tests; Grouping: Atherosclerotic vascular (Namely) (All in primary data.stw)<br>Group 1: Carotid, Brachiocephalic bifurcation<br>Group 2: Carotid, Brachiocephalic bifurcation |  |  |  |
| --- | --- | --- | --- | --- |
|  | t-value | df | p | Valid N<br>Carotid, Brachiocephalic<br>bifurcation |
| Max HR | -5.07001 | 12 | 0.000275 | 12 |
| Reached % | -5.00824 | 12 | 0.000305 | 12 |

3-H
| Variable | T-tests; Grouping: Atherosclerotic vascular (Namely) (All in primary data.stw)<br>Group 1: Carotid, Brachiocephalic bifurcation<br>Group 2: Carotid, Brachiocephalic bifurcation |  |
| --- | --- | --- |
|  | Valid N<br>Carotid, Brachiocephalic<br>bifurcation | Std.Dev.<br>Carotid, Brachiocephalic<br>bifurcation |
| Max HR | 2 | 10.210 |
| Reached % | 2 | 8.173 |

| Variable | T-tests; Grouping: Atherosclerotic vascular (Namely) (All in primary data.stw)<br>Group 1: Carotid, Brachiocephalic bifurcation<br>Group 2: Carotid, Brachiocephalic bifurcation |  |  |
| --- | --- | --- | --- |
|  | Std.Dev.<br>Carotid, Brachiocephalic<br>bifurcation | F-ratio<br>Variances | p<br>Variances |
| Max HR | 21.2132 | 4.317 | 0.123886 |
| Reached % | 15.0526 | 3.392 | 0.185215 |

3-I
| Variable | T-tests; Grouping: Carotid (All in primary data.stw)<br>Group 1: No<br>Group 2: Yes |  |  |  |  |  |
| --- | --- | --- | --- | --- | --- | --- |
|  | Mean<br>No | Mean<br>Yes | t-value | df | p | Valid N<br>No |
| Age | 53.33 | 66.08 | -4.77434 | 32 | 0.000038 | 20 |

| Variable | T-tests; Grouping: Carotid (All in primary data.stw) |  |  |  |  |  |
| --- | --- | --- | --- | --- | --- | --- |
|  | Group 1: No<br>Group 2: Yes |  |  |  |  |  |
|  | Mean<br>No | Mean<br>Yes | t-value | df | p | Valid N<br>No |
| Goal heart rate | 166.67 | 153.92 | 4.77434 | 32 | 0.000038 | 20 |
| METs | 7.35 | 5.56 | 2.60444 | 32 | 0.013842 | 20 |

| Variable | T-tests; Grouping: Carotid (All in primary data.stw) |  |  |  |  |
| --- | --- | --- | --- | --- | --- |
|  | Group 1: No<br>Group 2: Yes |  |  |  |  |
|  | Valid N<br>Yes | Std.Dev.<br>No | Std.Dev.<br>Yes | F-ratio<br>Variances | p<br>Variances |
| Age | 14 | 7.532 | 7.845 | 1.08495 | 0.849210 |
| Goal heart rate | 14 | 7.532 | 7.845 | 1.08495 | 0.849210 |
| METs | 14 | 2.508 | 0.597 | 17.65041 | 0.000005 |

3-J
| Variable | T-tests; Grouping: Brachiocephalic (All in primary data.stw) |  |  |  |  |  |
| --- | --- | --- | --- | --- | --- | --- |
|  | Group 1: No<br>Group 2: Yes |  |  |  |  |  |
|  | Mean<br>No | Mean<br>Yes | t-value | df | p | Valid N<br>No |
| Age | 54.18 | 66.65 | -4.38832 | 32 | 0.000116 | 22 |
| Goal heart rate | 165.82 | 153.35 | 4.38832 | 32 | 0.000116 | 22 |
| METs | 7.18 | 5.56 | 2.24411 | 32 | 0.031872 | 22 |

| Variable | T-tests; Grouping: Brachiocephalic (All in primary data.stw) |  |  |  |  |
| --- | --- | --- | --- | --- | --- |
|  | Group 1: No<br>Group 2: Yes |  |  |  |  |
|  | Valid N<br>Yes | Std.Dev.<br>No | Std.Dev.<br>Yes | F-ratio<br>Variances | p<br>Variances |
| Age | 12 | 8.165 | 7.432 | 1.20683 | 0.769942 |
| Goal heart rate | 12 | 8.165 | 7.432 | 1.20683 | 0.769942 |
| METs | 12 | 2.452 | 0.587 | 17.45301 | 0.000023 |

3-K
| Variable | T-tests; Grouping: Arterial Hypertension (All in primary data.stw) |  |  |  |  |  |
| --- | --- | --- | --- | --- | --- | --- |
|  | Group 1: Yes<br>Group 2: No |  |  |  |  |  |
|  | Mean<br>Yes | Mean<br>No | t-value | df | p | Valid N<br>Yes |
| Age | 62.13 | 54.62 | 2.55288 | 36 | 0.015067 | 21 |
| SBP rest | 126.00 | 118.12 | 2.03149 | 36 | 0.049637 | 21 |
| Goal heart rate | 157.87 | 165.38 | -2.55288 | 36 | 0.015067 | 21 |
| METs | 5.90 | 7.44 | -2.45597 | 36 | 0.019008 | 21 |
| eGFR (2021 CKD-EPI Creatinine) | 77.32 | 87.88 | -2.16746 | 36 | 0.036892 | 21 |

| Variable | T-tests; Grouping: Arterial Hypertension (All in primary data.stw) |  |  |  |  |
| --- | --- | --- | --- | --- | --- |
|  | Group 1: Yes<br>Group 2: No |  |  |  |  |
|  | Valid N<br>No | Std.Dev.<br>Yes | Std.Dev.<br>No | F-ratio<br>Variances | p<br>Variances |
| Age | 17 | 8.924 | 9.128 | 1.04632 | 0.910887 |
| SBP rest | 17 | 12.235 | 11.450 | 1.14179 | 0.797035 |
| Goal heart rate | 17 | 8.924 | 9.128 | 1.04632 | 0.910887 |
| METs | 17 | 1.407 | 2.395 | 2.89929 | 0.026188 |

| Variable | T-tests; Grouping: Arterial Hypertension (All in primary data.stw)<br>Group 1: Yes<br>Group 2: No |  |  |  |  |
| --- | --- | --- | --- | --- | --- |
|  | Valid N<br>No | Std.Dev.<br>Yes | Std.Dev.<br>No | F-ratio<br>Variances | p<br>Variances |
| eGFR (2021 CKD-EPI Creatinine) | 17 | 15.615 | 14.046 | 1.23599 | 0.674001 |

3-L
| Variable | T-tests; Grouping: Reaction type ( positive/negative) (All in primary data.stw)<br>Group 1: Positive<br>Group 2: Negative |  |  |  |  |  |
| --- | --- | --- | --- | --- | --- | --- |
|  | Mean<br>Positive | Mean<br>Negative | t-value | df | p | Valid N<br>Positive |
| WT | 106.25 | 144.12 | -2.18459 | 27 | 0.037776 | 12 |

| Variable | T-tests; Grouping: Reaction type ( positive/negative) (All in primary data.stw)<br>Group 1: Positive<br>Group 2: Negative |  |  |  |  |
| --- | --- | --- | --- | --- | --- |
|  | Valid N<br>Negative | Std.Dev.<br>Positive | Std.Dev.<br>Negative | F-ratio<br>Variances | p<br>Variances |
| WT | 17 | 37.119 | 51.181 | 1.901152 | 0.283098 |

3-M
| Variable | T-tests; Grouping: Reason of discontinuation (All in primary data.stw)<br>Group 1: Horizontal ST depression >1mm<br>Group 2: Reach goal HR |  |  |  |
| --- | --- | --- | --- | --- |
|  | Mean<br>Horizontal ST depression<br>>1mm | Mean<br>Reach goal HR | t-value | df |
| Creatinine (μmol/L) | 98.62 | 82.62 | 2.39629 | 36 |

| Variable | T-tests; Grouping: Reason of discontinuation (All in primary data.stw)<br>Group 1: Horizontal ST depression >1mm<br>Group 2: Reach goal HR |  |  |
| --- | --- | --- | --- |
|  | p | Valid N<br>Horizontal ST depression >1mm | Valid N<br>Reach goal HR |
| Creatinine (μmol/L) | 0.021881 | 6 | 32 |
|  | Std.Dev.<br>Horizontal ST depression<br>>1mm | Std.Dev.<br>Reach goal HR | F-ratio<br>Variances |
| Creatinine (μmol/L) | 23.934 | 13.007 | 3.385970 |

| Variable | T-tests; Grouping: Reason of discontinuation (All in primary data.stw)<br>Group 1: Horizontal ST depression >1mm<br>Group 2: Reach goal HR |
| --- | --- |

|  | p<br>Variances |
| --- | --- |
| Creatinine (μmol/L) | 0.029651 |

Interestingly, the sensitivity and specificity of the bicycle ergometry showed a very low level and not sufficient for confirming the diagnosis of ischemic heart disease. (*Table 4*)

**Table 4:** The specificity and sensitivity of the bicycle ergometry. (*) These values are dependent on disease prevalence.

| Statistic | Value | 95% CI |
| --- | --- | --- |
| Sensitivity | 57.89% | 33.50% to 79.75% |
| Specificity | 61.11% | 35.75% to 82.70% |
| Positive Likelihood Ratio | 1.49 | 0.74 to 2.98 |
| Negative Likelihood Ratio | 0.69 | 0.36 to 1.31 |
| Disease prevalence (*) | 51.35% | 34.40% to 68.08% |
| Positive Predictive Value (*) | 61.11% | 43.96% to 75.89% |
| Negative Predictive Value (*) | 57.89% | 41.95% to 72.35% |
| Accuracy (*) | 59.46% | 42.10% to 75.25% |

All the correlation results between the continuous variables presented in supplementary material. (Supplementary file 1

## Dissuasion

Interestingly, on the CTP test, 19 (50 %) participants with coronary artery atherosclerosis, 5 (26 %) of them with significant atherosclerosis (stenosis >60 %). Furthermore, out of the 19 patients with significant atherosclerosis of the coronary artery, 11 (57 %) had myocardial perfusion defect after stress test with ATP.

Out of the 11 patients with positive myocardial perfusion defect after the stress test, 4 (36 %) participants with positive bicycle ergometry test.

More interesting findings, in the whole sample there is 19 (50 %) participants with positive myocardial perfusion defect, and only 5 (26 %) of them with significant coronary artery stenosis. Out of the 19 positive myocardial perfusion defect, 11 (57 %) with positive/suspected results on the bicycle ergometry.

2 (5.26 %) participants out of the 38 with positive results on the bicycle ergometry with heart rate < 85 % of the required goal heart rate (220-age). In other participants with positive (11)/suspected (8) results on the bicycle ergometry with heart rate > 85 % of the required heart rate goal.

In the whole sample there is 12 (31.58 %) positive, 9 (23.68 %) suspected, and 17 (44.47 %) negative on the bicycle ergometry. However, on the CTP, out of the 12 positive bicycle ergometry, only 5 (41.67 %) with positive myocardial perfusion defect after stress test with ATP. Moreover, out of the 9 suspected participants, 6 (66.67 %) had positive myocardial perfusion defect after stress test with ATP.

The presented results showed that bicycle ergometry suffers from severe limitations in terms of specificity and sensitivity in the diagnosis of IHD. Additional methods are required to improve the diagnostic accuracy of the bicycle ergometry such as the exhaled breath analysis and single channel electrocardiography ^[11]^.

## Conclusions

On the light of the presented results, the diagnosis of IHD using bicycle ergometry independently is ultra extremely not sufficient and further investigations required to improve the diagnostic accuracy of the bicycle ergometry. As well as to innovate novel methods for early detection of IHD through the application of the current advances in technologies such as the use of artificial intelligence (machine learning models) in the interpretation of the electrocardiography monitoring for patient at home bases using their watches or phones ^[6, 12]^. Additionally, supplementary methods can be combined with the bicycle ergometry to enhance it is diagnostic accuracy.

## Supporting information

Supp. File 1. correlations

## Data Availability

All data produced in the present work are contained in the manuscript

## List of abbreviations

IHD: Ischemic heart disease
CTP: stress computed tomography myocardial perfusion imaging

## Decelerations

### Ethics approval and consent to participate

The study approved by the Sechenov University, Russia, from “Ethics Committee Requirement № 19-23 from 26.10.2023”.

The study is under consideration by the clinicaltrails.gov for registration.

### Consent for publication

Written informed consent was obtained from the participants for publication of study results and any accompanying images.

### Availability of data and materials

applicable on reasonable request.

### Funding’s

The work of Philipp Kopylov and Peter Chomakhidze was financed by the government assignment 1023022600020-6 «Application of mass spectrometry and exhaled air emission spectrometry for cardiovascular risk stratification».

### Authors’ contributions

**Basheer Abdullah Marzoog**, is the writer, researcher, collected and analyzed data, and revised the manuscript, **Peter Chomakhidze and Philipp Kopylov** revised the manuscript. All authors have read and approved the manuscript.

## Acknowledgments

not applicable

## Authors’ information

**Basheer Abdullah Marzoog**, World-Class Research Center «Digital Biodesign and Personalized Healthcare», I.M. Sechenov First Moscow State Medical University (Sechenov University), 119991 Moscow, Russia; postal address: Russia, Moscow, 8-2 Trubetskaya street, 119991. (, +79969602820). Scopus ID: 57486338800. **Peter Chomakhidze**, World-Class Research Center «Digital Biodesign and Personalized Healthcare», I.M. Sechenov First Moscow State Medical University (Sechenov University), 119991 Moscow, Russia; postal address: Russia, Moscow, 8-2 Trubetskaya street, 119991. **Philipp Kopylov**, World-Class Research Center «Digital Biodesign and Personalized Healthcare», I.M. Sechenov First Moscow State Medical University (Sechenov University), 119991 Moscow, Russia; postal address: Russia, Moscow, 8-2 Trubetskaya street, 119991.

The paper has not been submitted elsewhere

